# Effect of phone text message reminders on compliance with rabies post-exposure prophylaxis following dog-bites in rural Kenya

**DOI:** 10.1101/2022.06.16.22276500

**Authors:** Veronicah Mbaire Chuchu, Mutono Nyamai, Philet Bichanga, Kitala Philip, Daniel Ksee, Mathew Muturi, Athman Mwatondo, Caroline Nasimiyu, Lawrence Akunga, Amine Amiche, Katie Hampson, SM Thumbi

## Abstract

**Context:** Prompt administration of post exposure prophylaxis (PEP) is one of the key strategies for ending human deaths from rabies. Delay in seeking first dose of rabies PEP or failure to complete the recommended PEP dosage may result in clinical rabies and death.

**Objectives:** To assess the efficacy of short message system (SMS) phone texts on adherence to scheduled PEP doses among bite patients in rural eastern Kenya.

**Trial design and methods:** We conducted a single arm before-after field trial that compared adherence among bite patients presenting at Makueni Referral Hospital in October – December 2018 (control group) and January – March 2019 (intervention group that received an SMS reminder one day prior to their scheduled PEP doses). Data on demographics, socio-economic status, circumstances surrounding the bite, expenditures related to the bite were collected for all patients.

**Results:** A total of 186 bite patients were enrolled in the study, with 82 (44%) being in the intervention and 104 (56%) in the control group. The odds of PEP completion was three times (OR 3.37, 95% CI 1.28, 10.20) among patients that received the SMS reminder compared to those that did not. The intervention group had better compliance on the scheduled doses 2 to 5 with a mean deviation of 0.18 days compared to 0.79 days for the control group (*p* = 0.004). The main reasons for non-compliance included lack of funds (30%), forgetfulness (23%) on days for follow-up treatment, among others. Although the majority of bite patients (94%) were under the Makueni medical insurance cover and did not pay for PEP, nearly all (96%, n=179) the bite patients incurred indirect costs of transport at an average of 4 USD (0 - 45 USD) per visit.

**Conclusion:** This study suggests integrating SMS reminders in healthcare service delivery increases compliance to PEP and may strengthen rabies control and elimination strategies.

**Trial registration:** The study trial is registered at US National Institute of Health (clinicalTrial.gov) identifier number NCT05350735. https://clinicaltrials.gov/ct2/show/NCT05350735

## Introduction

Rabies, a fatal viral disease transmitted to humans mainly by domestic dogs, is a neglected zoonosis that primarily affects underserved populations that have limited access to health care. Every year, rabies is estimated to kill 59,000 people globally, mostly children 15 years and below in Africa and Asia [1, 2]. This is despite the development of effective vaccines against rabies in humans, and in dogs [3]. Although rabies is always fatal once clinical signs manifest, the disease is preventable with timely treatment after exposure to the rabies virus. The World Health Organization (WHO) recommendations for bite patients is immediate thorough cleaning of the wound with soap and water or virucidal agents for approximately 15 minutes, followed by administration of anti-rabies vaccine, and where there are multiple severe bites particularly to the head and upper trunk, infiltration of rabies immunoglobulin into and around the wound(s) [4]. In developing countries, including Kenya, access to post-exposure vaccines is poor due to its unaffordable cost and unavailability [2]. Lack of access to post-exposure prophylaxis (PEP) or deviations from WHO recommendations such as delays in seeking PEP and incomplete courses of PEP increases the risk of clinical rabies and death [5,6]. To increase access and availability, WHO has updated their recommendations for rabies post-exposure vaccination regimens from the 0.5ml or 1 ml per dose of intramuscular (IM) Essen regimen schedule of five doses on day 0, 3, 7, 14 and 28 (5-dose Essen regimen) to either a one-week intradermal (ID) schedule that is dose sparing and cost-effective which consists of 0.1 ml of vaccine per dose given as two ID injections on day 0, 3 and 7 or a two week intramuscular (IM) injection 4-dose post-exposure prophylaxis regimen with injections on day 0, 3, 7 and between day 14 – 28 (4-dose Essen regimen) [4,5]. However, the ID regimen is partially implemented in many rabies endemic countries including Kenya which is still following the 5-dose IM Essen regimen.

In Kenya, rabies is endemic across the country and has been estimated to cause over 500 (95% CI 134, 1100) deaths annually [7]. A national strategic plan for the elimination of dog-mediated human rabies by 2030 was developed and adopted in 2014 [7]. The key components of the plan include mass dog vaccination, timely provision of pre- and post-exposure vaccines, enhanced rabies surveillance for both human and animal populations, and public health education and awareness on rabies, it’s prevention and control [7]. Additionally, the elimination activities would be phased starting with pilot counties selected for their high burden of rabies before scale-up to the rest of the country. Makueni County, where this study was conduted is one of the five Counties selected as pilot areas for Kenya’s rabies elimination strategy [8]. Previously, we have reported large variability in the availability of rabies post-exposure vaccines in the country with pilot counties having shorter stockout periods [9]. Information and medical products, vaccines and technolgies access and uptake are two of the six pillars of the WHO Health System Building Blocks. WHO encourages streghthening of health systems to achieve health goals set out [10]. Innovation such as the use of the mobile phone can play a part. The use of mobile phone applications in health has increased globally due to their availability and ability to deliver scalable interventions (11). Our previous studies have shown the role mobile phones can play in improving the detection of outbreaks of zoonotic diseases in the country [12,13]. Using text message reminders has been shown to improve patient compliance including for childhood immunization attendance and appointment reminders across different geographical settings and health care services [14,15,16]. The use of short message systems (SMS) has further been reported to improve participation by dog owners in mass dog vaccinations in Haiti [17], and compliance to PEP regimens in Tanzania [18]. In this study, our objective was to assess the effect of SMS reminders on compliance to the five-dose Essen rabies PEP regimen and the factors associated with compliance among dog-bite patients in Makueni County.

## Materials and methods

### Study area

The study was conducted in Makueni County, one of the five Counties selected as pilot areas for the implementation of the Kenya rabies elimination strategy. Makueni County is divided into six sub counties (Mbooni, Kaiti, Makueni, Kibwezi East, Kibwezi West, and Kilome) (Fig 1) and has an estimated human population of 987,653 as of 2019 [19]. The main referral hospital for the County is based at Wote town, which serves as the administrative centre of the County. Makueni County has a total of 248 public health facilities. Of these, 11 are subcounty health facilities and one county referral hospital. The 12 (sub-county and county referral) health facilities are mandated to provide anti-rabies vaccine to the community. However, other private health facilities provide the vaccines.

**Fig 1:**
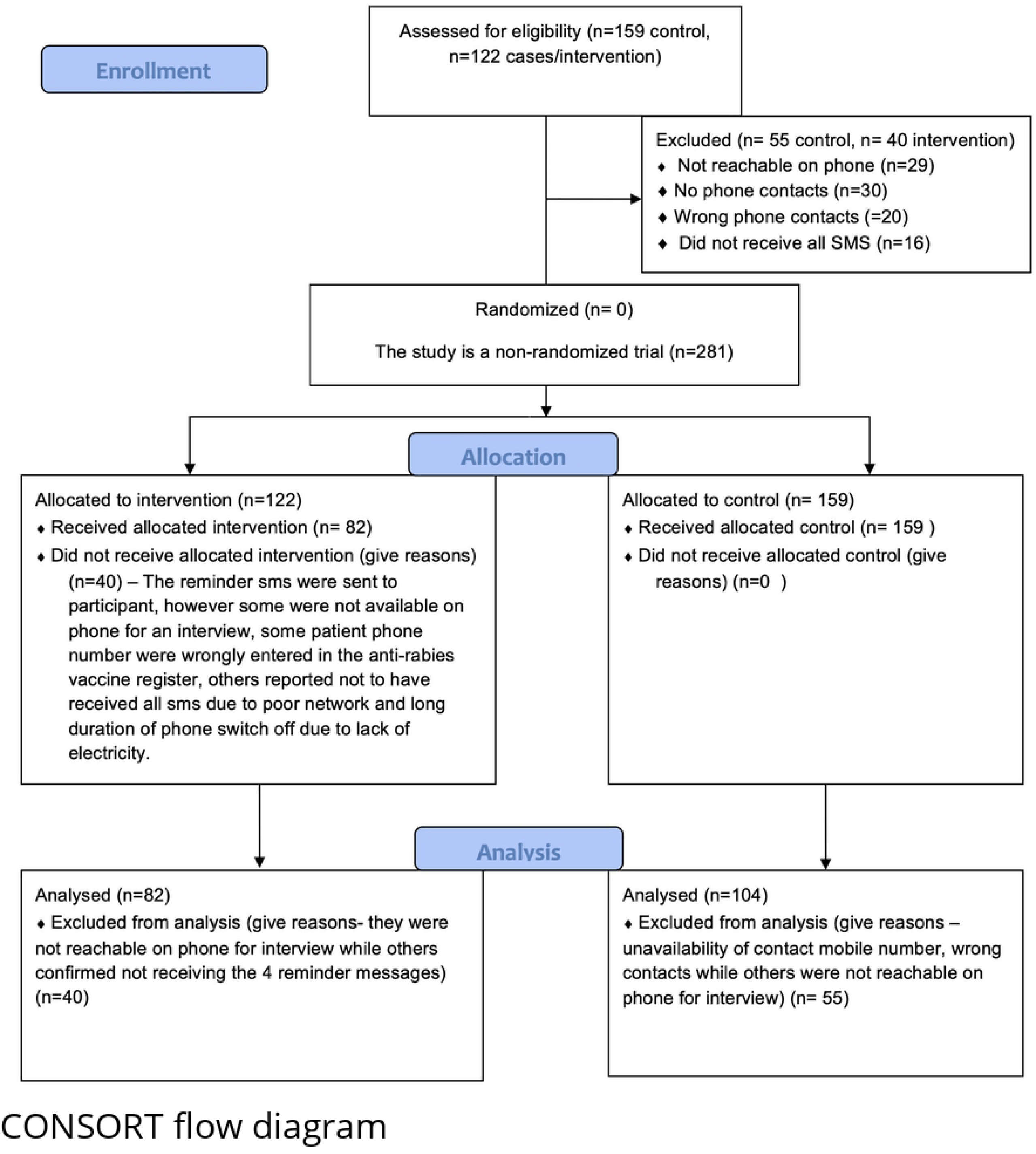
Flow diagram displaying the progress of all participants through the trial.

The cost of anti-rabies vaccine varies. In the public health facilities, the cost of the vaccine ranges from zero for patients under the Makueni universal health program, who are enrolled by paying an enrollment fee of USD 5, up to USD 8.5 per dose for patients not under the universal health program. In private health facilities, the cost of one dose may range from USD 10 to USD 25. The estimated annual bite incidence for Makueni county is 342 cases per 100,000 people per year [20].

### Study design and sample size calculation

We conducted a single arm before-after field trial among dog bite patients presenting at the Makueni County Referral hospital between October 2018 and March 2019. The study participants were allocated to either the control or the treatment group, based on recruitment time. Data on bite patient name, patients’ contacts, details of next of kin, site of bite, species of the biting animal, bite severity, vaccination status of the biting animal and date of each PEP dose received were extracted from the anti-rabies vaccine register at the County Referral Hospital. Beginning January 2019, an SMS reminder written in both English and the local dialect, Kamba, was sent out to all bite patients a day before their next dose of PEP. The SMS reminder was sent a day before each dose until the scheduled date of the last dose of PEP. Bite patients recruited into the study between October and December 2018 were designated as control group. As a routine, this group received a medical card indicating return date for the subsequent dose. Those recruited in January to March 2019 were designated as the intervention group. This group received the medical card and SMS reminders. To collect data on other factors affecting PEP completion and adherence, a phone interview was conducted to both groups in May, June and July 2019 with majority (76%) being contacted between May and June 2019. For the control group, all bite patients who responded to the phone interview were enrolled in the study, while in the intervention group, all bite patients who responded to the interview and confirmed to have received all four SMS reminders were considered for final data analysis.

The bite case data from the referral hospital available before the start of the study showed an average of 30 bite cases per month. To establish the number of human dog-bite cases to be recruited to either the control or the intervention group, we hypothesized SMS reminders would increase compliance and calculated the required sample size to detect a 20% increase in compliance in the group receiving SMS reminders compared to the group not receiving the reminders, with 80% power and at the significance level of 0.05. We estimated a sample size of 90 bite patients was required for each of the control and intervention groups assuming an increase in compliance from 60% to 80% following the introduction of the SMS reminder. The power calculations were carried out using the “pwr” package [22] of the R statistical computing software [23]. However, all dog bite patients recorded in the register during the study period and in possession of a phone were enrolled in the study to cater for withdrawals. The primary outcome measures were the number of participants that completed the five-doses Essen rabies vaccine regimen in the control versus the intervention group and the number of participants adhering to the scheduled date of the five-dose Essen vaccine regimen in the two groups.

### Data collection

For each study participant, a questionnaire was administered by phone call at the end of the PEP period, after providing verbal consent to participate in the study. For bite patient below 18 years, parent or guardian was interviewd. The data collected included demographics of the bite patient (age, sex), home location, characteristics of the bite, PEP compliance (date of bite and date of subsequent PEP injections) and data on putative factors affecting PEP completion which included the SMS reminder, ownership of health insurance cover, education level of bite patient, dog ownership status, vaccination status of the biting animal, fate of the biting dog (alive, dead or disappeared), time taken to reach the health facility, means of transport used, total cost of transport, if the bite patient was accompanied to the health facility, total cost of PEP, source of money incurred, household head occupation and age, total monthly household income, number of people in the household and livestock ownership status.

### Data analysis

The study questionnaires were programmed in the CommCare^®^ data collection tool to allow for electronic data capture using mobile phones. Data was then downloaded as a Microsoft Excel file, and statistical analysis was undertaken using the R computing language [23]. To assess the factors associated with completion of the five doses of PEP, the uptake of PEP was dichotomously classified based on whether the patient completed the full regimen or not (1 or 0) and univariate analysis was carried out on the different independent variables (Chi-square tests and t-tests for categorical and numerical variables respectively).

To understand the factors associated with compliance and completion of the anti-rabies vaccine schedule, multivariable logistic regressions were conducted for compliance of each dose. The akaike information criterion backward and forward stepwise algorithm was used to identify the suitable factors for each model.

### Ethical clearance

The study received ethical approval from Kenya Medical Research Institute/Scientific and Ethics Review Unit (SERU) (Ref No. KEMRI/SERU/CGHR/046/3268).

## Results

### Demographic and socio-economic characteristics

A total of 281 bite patients were recruited in the study, 159 (57%) in the control group and 122 (43%) in the intervention group. About a third (n=55, 35%) in the control group were excluded from the study due to unavailability of contact mobile numbers, wrong phone numbers while others were not reachable on phone for interview. In the intervention group, 40 (33%) bite patients we excluded due to their unavailability on phone for interview, wrong contact numbers while others reported not to have received all the reminders due to poor phone network and long duration of phone switch off due to lack of electricity.

A total of 186 bite patients were considered for analysis, 104 (56%) in the control group and 82 (44%) in the intervention group. More than half (n=101, 54%) of the bite patients were male, and the median age of the bite patients was 14 years (IQR 8, 38 years) (Table 1). The majority (n=104, 56%) of the bite patients were children below the age of 15 years (Fig 2). Half (n=93) of the patients were in primary school followed by 50 (27%) and 39 (21%) in secondary and tertiary school respectively. The majority (n=173, 93%) of the bite patients resided in rural areas of Makueni County. The average household size of the bite patients was six people (range 1-32). The main occupation for most households was farming, followed by business. Nearly two thirds (n=123, 66%) of the households had a monthly income of less than USD 100 (Table 1).

**Table 1:**
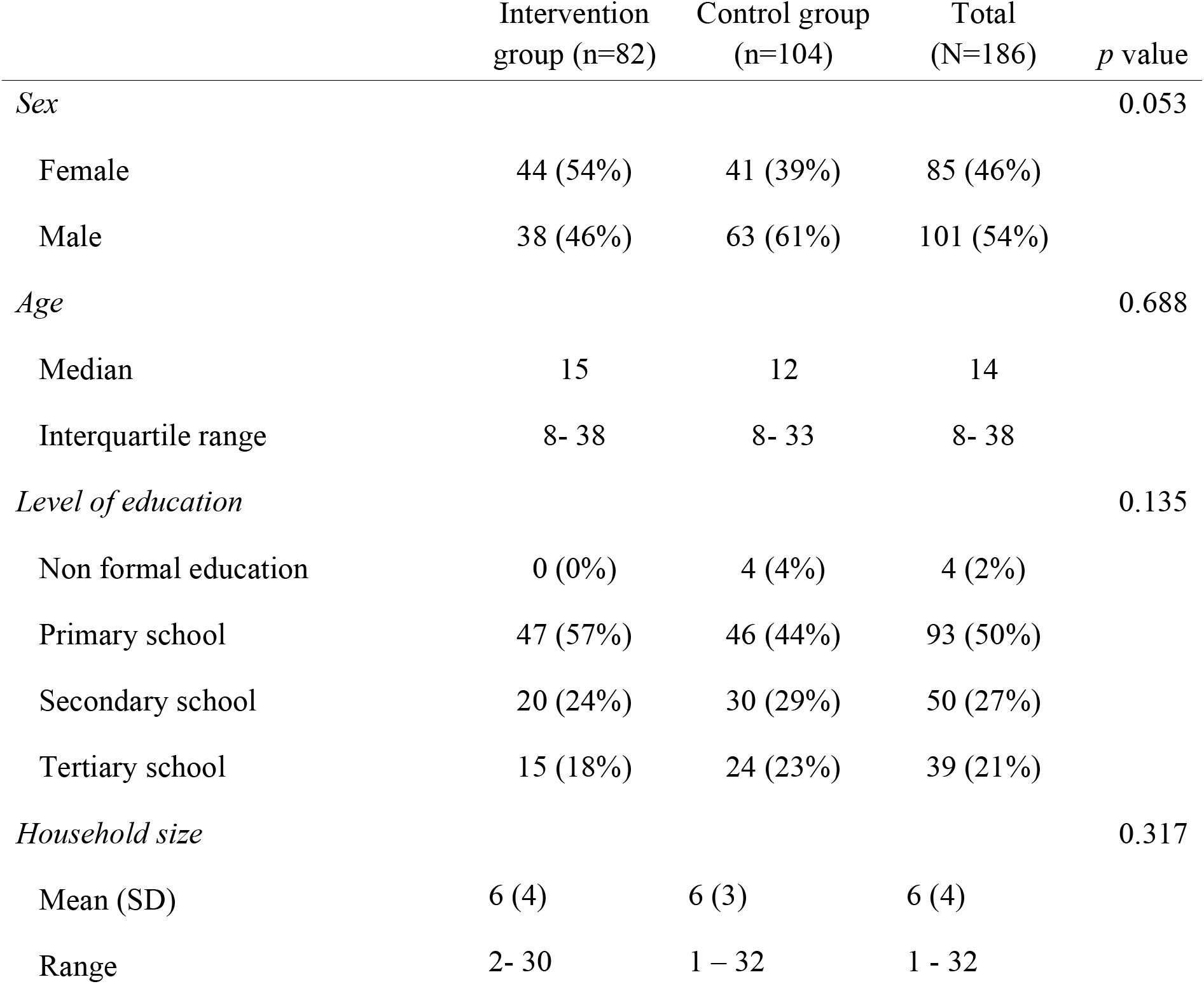
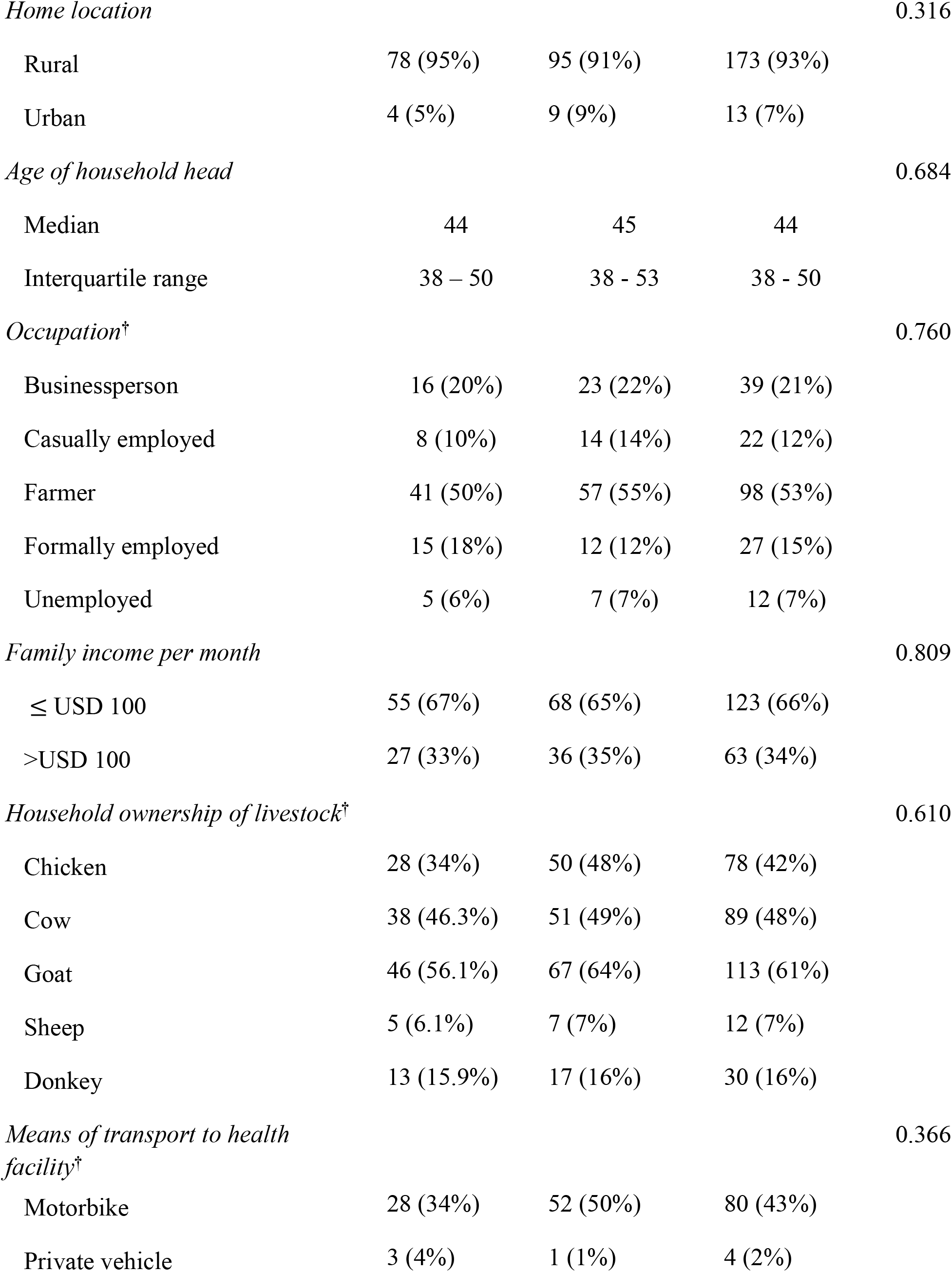
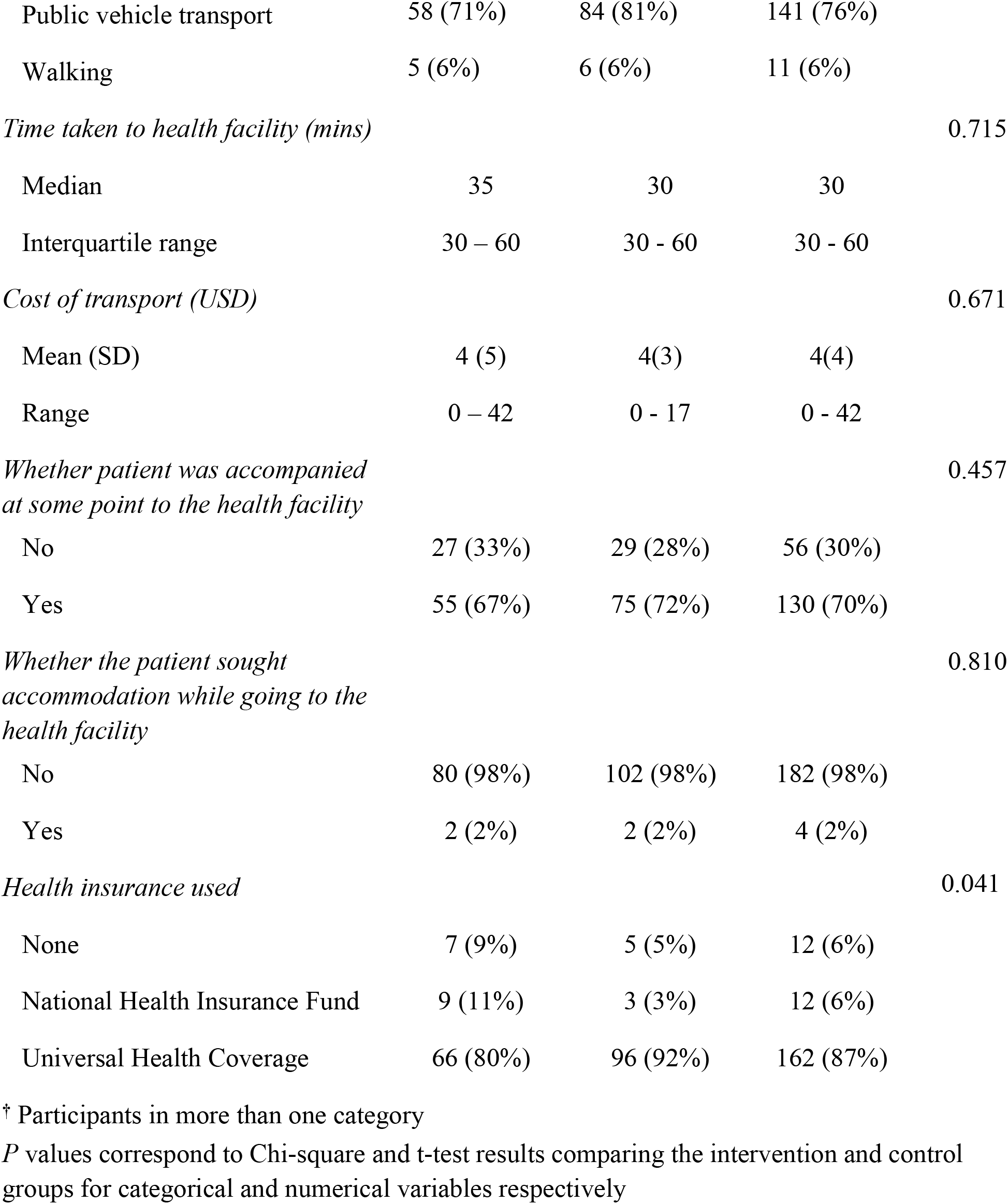
Demographics of the bite patients.

|  | Intervention group (n=82) | Control group (n=104) | Total (N=186) | *p* value |
| --- | --- | --- | --- | --- |
| *Sex* |  |  |  | 0.053 |
| Female | 44 (54%) | 41 (39%) | 85 (46%) |  |
| Male | 38 (46%) | 63 (61%) | 101 (54%) |  |
| *Age* |  |  |  | 0.688 |
| Median | 15 | 12 | 14 |  |
| Interquartile range | 8- 38 | 8- 33 | 8- 38 |  |
| *Level of education* |  |  |  | 0.135 |
| Non formal education | 0 (0%) | 4 (4%) | 4 (2%) |  |
| Primary school | 47 (57%) | 46 (44%) | 93 (50%) |  |
| Secondary school | 20 (24%) | 30 (29%) | 50 (27%) |  |
| Tertiary school | 15 (18%) | 24 (23%) | 39 (21%) |  |
| *Household size* |  |  |  | 0.317 |
| Mean (SD) | 6 (4) | 6 (3) | 6 (4) |  |
| Range | 2- 30 | 1 – 32 | 1 - 32 |  |
| *Home location* |  |  |  | 0.316 |
| Rural | 78 (95%) | 95 (91%) | 173 (93%) |  |
| Urban | 4 (5%) | 9 (9%) | 13 (7%) |  |
| *Age of household head* |  |  |  | 0.684 |
| Median | 44 | 45 | 44 |  |
| Interquartile range | 38 – 50 | 38 - 53 | 38 - 50 |  |
| *Occupation<sup>†</sup>* |  |  |  | 0.760 |
| Businessperson | 16 (20%) | 23 (22%) | 39 (21%) |  |
| Casually employed | 8 (10%) | 14 (14%) | 22 (12%) |  |
| Farmer | 41 (50%) | 57 (55%) | 98 (53%) |  |
| Formally employed | 15 (18%) | 12 (12%) | 27 (15%) |  |
| Unemployed | 5 (6%) | 7 (7%) | 12 (7%) |  |
| *Family income per month* |  |  |  | 0.809 |
| ≤ USD 100 | 55 (67%) | 68 (65%) | 123 (66%) |  |
| >USD 100 | 27 (33%) | 36 (35%) | 63 (34%) |  |
| *Household ownership of livestock<sup>†</sup>* |  |  |  | 0.610 |
| Chicken | 28 (34%) | 50 (48%) | 78 (42%) |  |
| Cow | 38 (46.3%) | 51 (49%) | 89 (48%) |  |
| Goat | 46 (56.1%) | 67 (64%) | 113 (61%) |  |
| Sheep | 5 (6.1%) | 7 (7%) | 12 (7%) |  |
| Donkey | 13 (15.9%) | 17 (16%) | 30 (16%) |  |
| *Means of transport to health facility<sup>†</sup>* |  |  |  | 0.366 |
| Motorbike | 28 (34%) | 52 (50%) | 80 (43%) |  |
| Private vehicle | 3 (4%) | 1 (1%) | 4 (2%) |  |
| Public vehicle transport | 58 (71%) | 84 (81%) | 141 (76%) |  |
| Walking | 5 (6%) | 6 (6%) | 11 (6%) |  |
| *Time taken to health facility (mins)* |  |  |  | 0.715 |
| Median | 35 | 30 | 30 |  |
| Interquartile range | 30 – 60 | 30 - 60 | 30 - 60 |  |
| *Cost of transport (USD)* |  |  |  | 0.671 |
| Mean (SD) | 4 (5) | 4(3) | 4(4) |  |
| Range | 0 – 42 | 0 - 17 | 0 - 42 |  |
| *Whether patient was accompanied at some point to the health facility* |  |  |  | 0.457 |
| No | 27 (33%) | 29 (28%) | 56 (30%) |  |
| Yes | 55 (67%) | 75 (72%) | 130 (70%) |  |
| *Whether the patient sought accommodation while going to the health facility* |  |  |  | 0.810 |
| No | 80 (98%) | 102 (98%) | 182 (98%) |  |
| Yes | 2 (2%) | 2 (2%) | 4 (2%) |  |
| *Health insurance used* |  |  |  | 0.041 |
| None | 7 (9%) | 5 (5%) | 12 (6%) |  |
| National Health Insurance Fund | 9 (11%) | 3 (3%) | 12 (6%) |  |
| Universal Health Coverage | 66 (80%) | 96 (92%) | 162 (87%) |  |

† Participants in more than one category
*P* values correspond to Chi-square and t-test results comparing the intervention and control groups for categorical and numerical variables respectively

**Fig 2:**
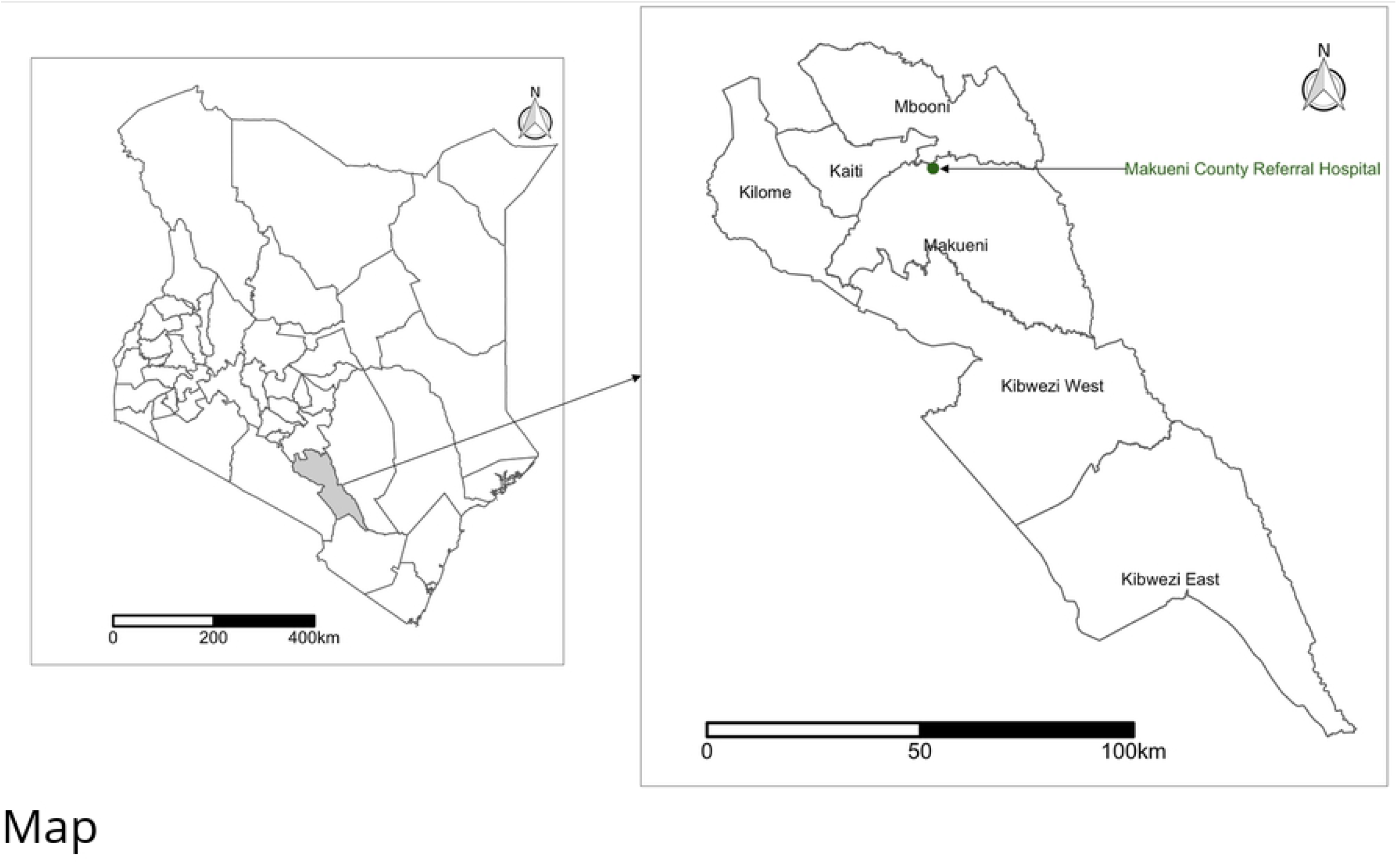
Map of Kenya highlighting the location of Makueni County (left) and the Makueni County Referral hospital in Makueni subcounty (right). Shapefile source: Database of Global Administrative Areas [21].

In total, 141 bite patients (76%) used public transportation to reach the health facilities. The average cost of transportation spent by the bite patients was USD 4 per visit. The average time taken to reach the health facility was 47 minutes (range 2 - 240 minutes). Seventy percent (n=130) of the bite patients were accompanied to the health facility at some point during the treatment period while 2% (n=4) of the patients sought accommodation while going to the health facility. Almost all (n=174, 94%) of the bite patients were beneficiaries of a health insurance scheme which reduced the cost of treatment at the health facility (Table 1).

### Characteristics of the bite and biting animal

The most common bite sites were to the legs (n=110, 59%) and the arms (n=56, 30%). Most of the bites were classified as either category two (n=88, 47%) and category three (n=81, 44%) as per WHO categorization of bite wounds [5]. More than half (n=109, 59%) of the patient were bitten by dogs known to them but not their own dogs whereas nearly a third (n=57, 31%) were bitten by their own dogs. Only 43% of the biting dogs (n=80) had a history of vaccination, while 69% (n=128) were alive, and 15% (n=27) dead at the time of the interview (Table 2).

**Table 2:**
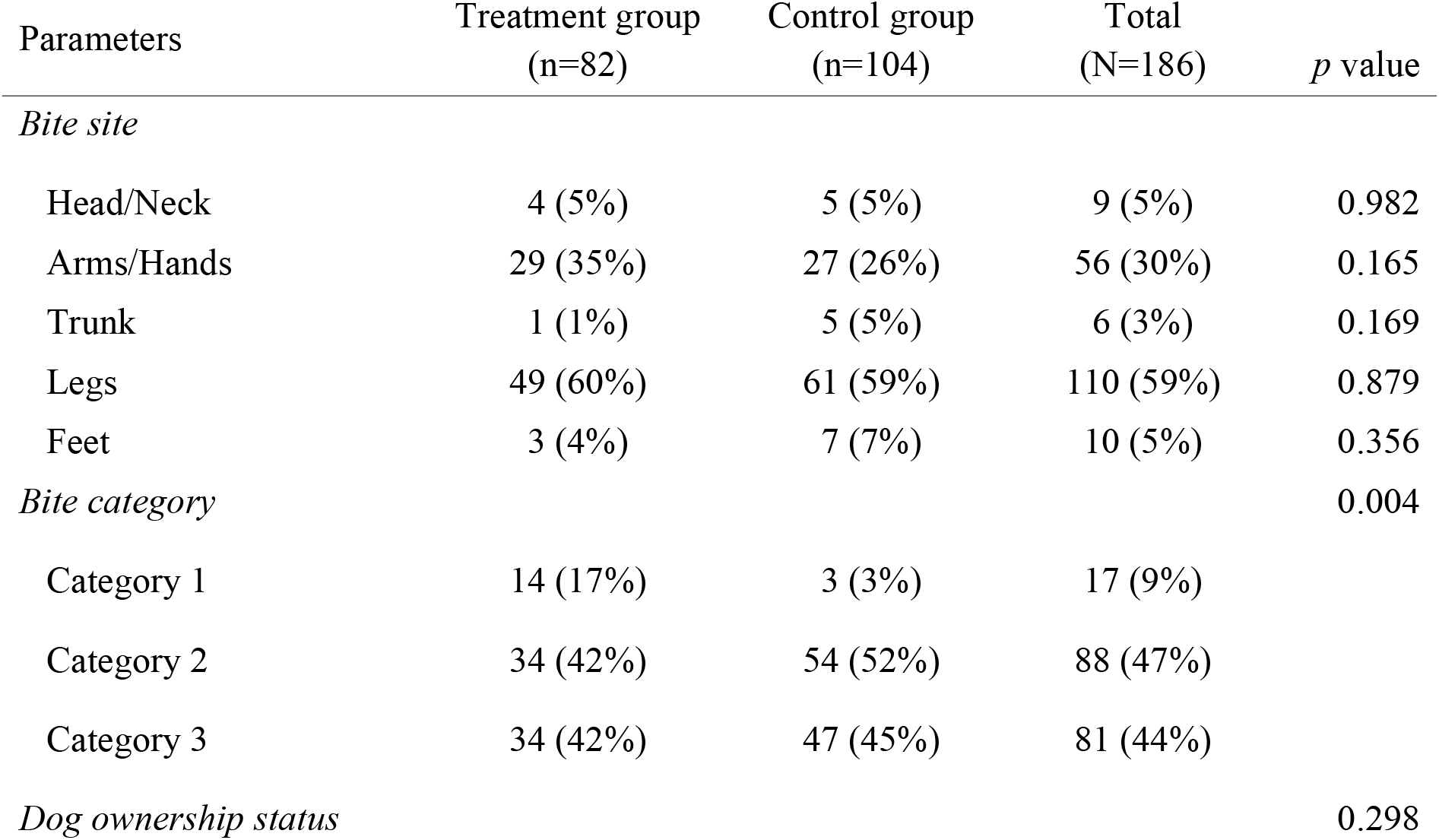
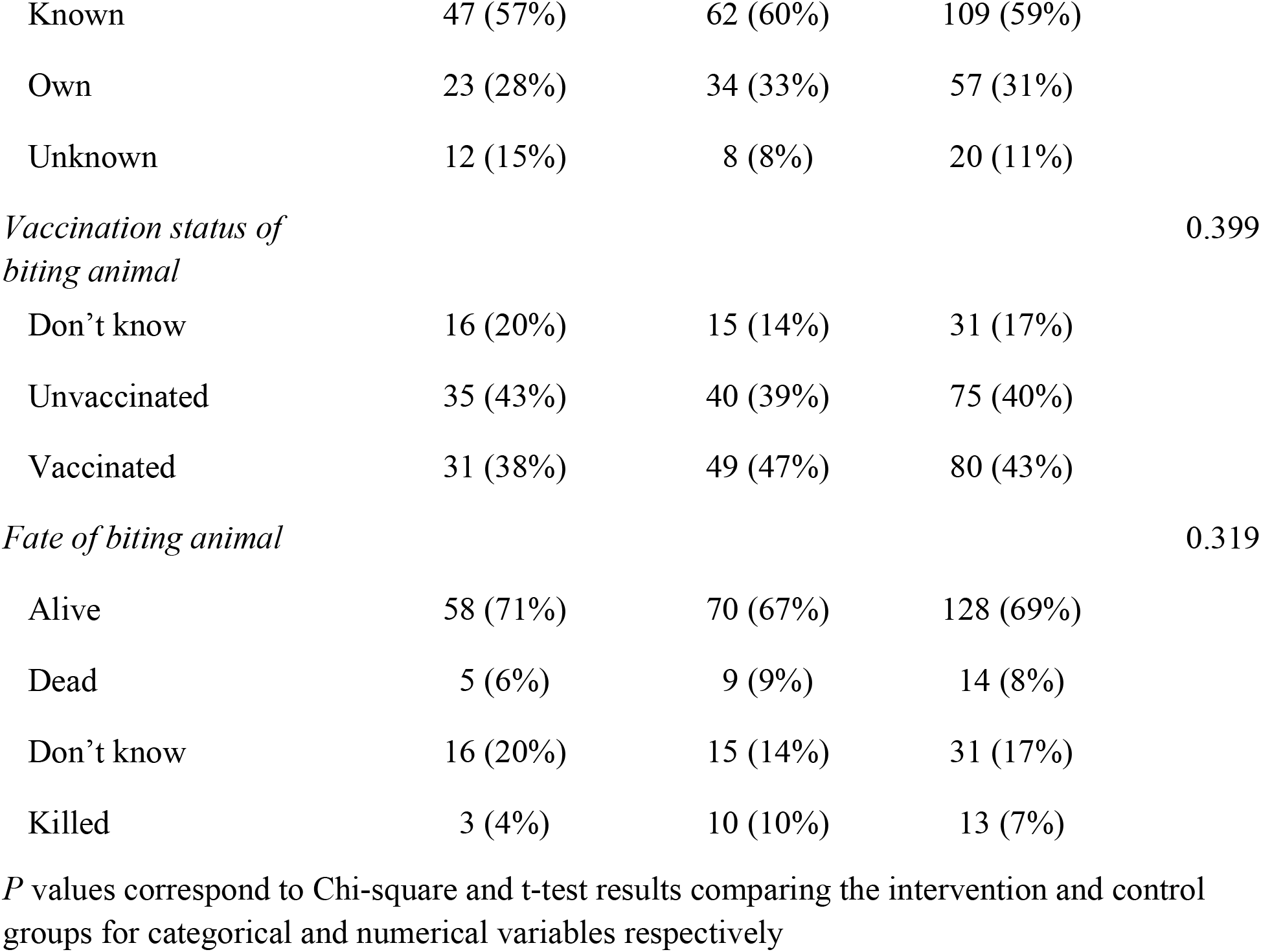
Characteristics of bites and the status of the biting animal.

| Parameters | Treatment group<br>(n=82) | Control group<br>(n=104) | Total<br>(N=186) | *p* value |
| --- | --- | --- | --- | --- |
| *Bite site* |  |  |  |  |
| Head/Neck | 4 (5%) | 5 (5%) | 9 (5%) | 0.982 |
| Arms/Hands | 29 (35%) | 27 (26%) | 56 (30%) | 0.165 |
| Trunk | 1 (1%) | 5 (5%) | 6 (3%) | 0.169 |
| Legs | 49 (60%) | 61 (59%) | 110 (59%) | 0.879 |
| Feet | 3 (4%) | 7 (7%) | 10 (5%) | 0.356 |
| *Bite category* |  |  |  | 0.004 |
| Category 1 | 14 (17%) | 3 (3%) | 17 (9%) |  |
| Category 2 | 34 (42%) | 54 (52%) | 88 (47%) |  |
| Category 3 | 34 (42%) | 47 (45%) | 81 (44%) |  |
| *Dog ownership status* |  |  |  | 0.298 |
| Known | 47 (57%) | 62 (60%) | 109 (59%) |  |
| Own | 23 (28%) | 34 (33%) | 57 (31%) |  |
| Unknown | 12 (15%) | 8 (8%) | 20 (11%) |  |
| *Vaccination status of biting animal* |  |  |  | 0.399 |
| Don't know | 16 (20%) | 15 (14%) | 31 (17%) |  |
| Unvaccinated | 35 (43%) | 40 (39%) | 75 (40%) |  |
| Vaccinated | 31 (38%) | 49 (47%) | 80 (43%) |  |
| *Fate of biting animal* |  |  |  | 0.319 |
| Alive | 58 (71%) | 70 (67%) | 128 (69%) |  |
| Dead | 5 (6%) | 9 (9%) | 14 (8%) |  |
| Don't know | 16 (20%) | 15 (14%) | 31 (17%) |  |
| Killed | 3 (4%) | 10 (10%) | 13 (7%) |  |

*P* values correspond to Chi-square and t-test results comparing the intervention and control groups for categorical and numerical variables respectively

### Effect of SMS reminders on compliance to the PEP regimen

Among the 104 participants recruited into the control group, 81 (78%) completed the five doses of the PEP vaccine. For the 82 participants in the intervention group, 76 (93%) completed the PEP doses resulting in a 15% increase in completion rates. Out of the 29 (16%) bite patients who did not complete the five doses of PEP, 23 (n=79%) were in the control period.

The odds of PEP completion with SMS reminders was three times (OR 3.37, 95% CI 1.28, 10.20) in the intervention group compared to the participants in the control group (Table 3). We studied the compliance in the day of PEP administration for each of the five doses against the WHO recommendation for the Essen regimen for day 0, 3, 7, 14 and 28 days. Nearly a third (n=59, 32%) of the bite patients received the first dose of PEP less than 24 hours after the bite while 45% took the dose one to two days after the bite. Compliance to the scheduled date of PEP improved to more than 70% receiving second, third, fourth and fifth dose on day 3, 7, 14 and 28 after the bite respectively. The average time from bite to first dose of PEP was 1.99 days with no statistical differences between control and intervention groups. However, we found the intervention group had better compliance on the scheduled doses 2 to 5 with a mean deviation of 0.18 days compared to 0.79 days for the control group (*p* = 0.004) (Fig 3).

**Table 3:**
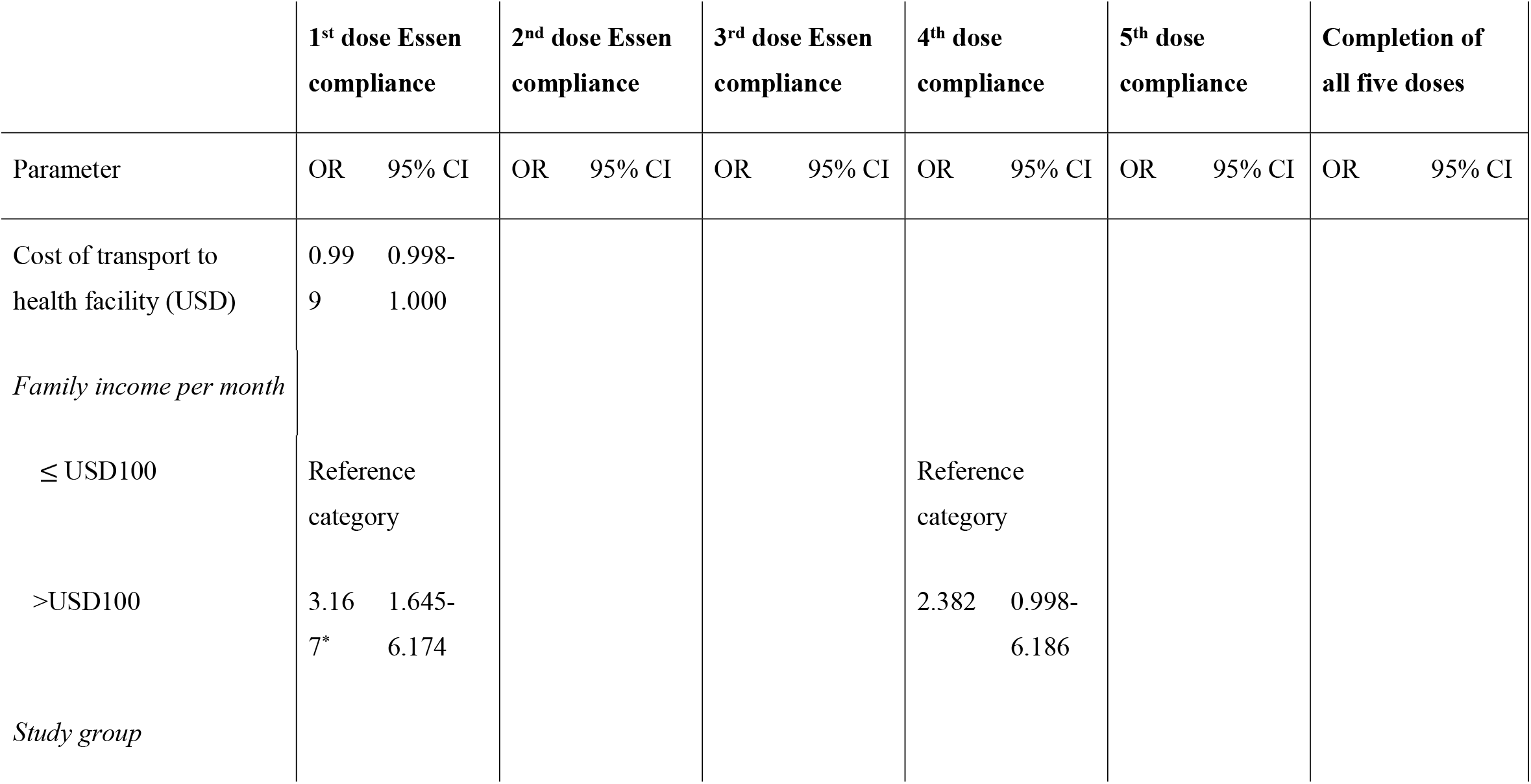
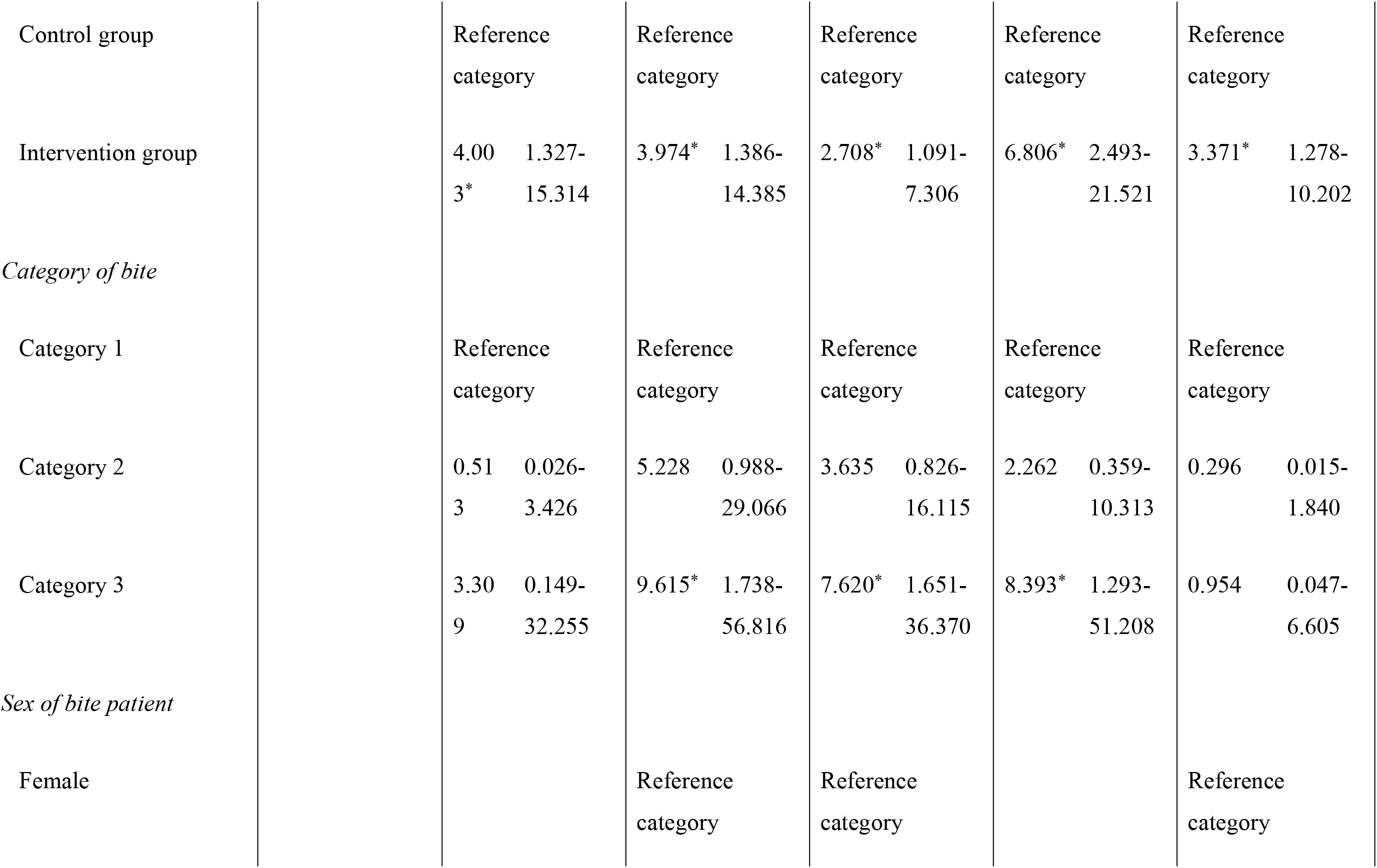
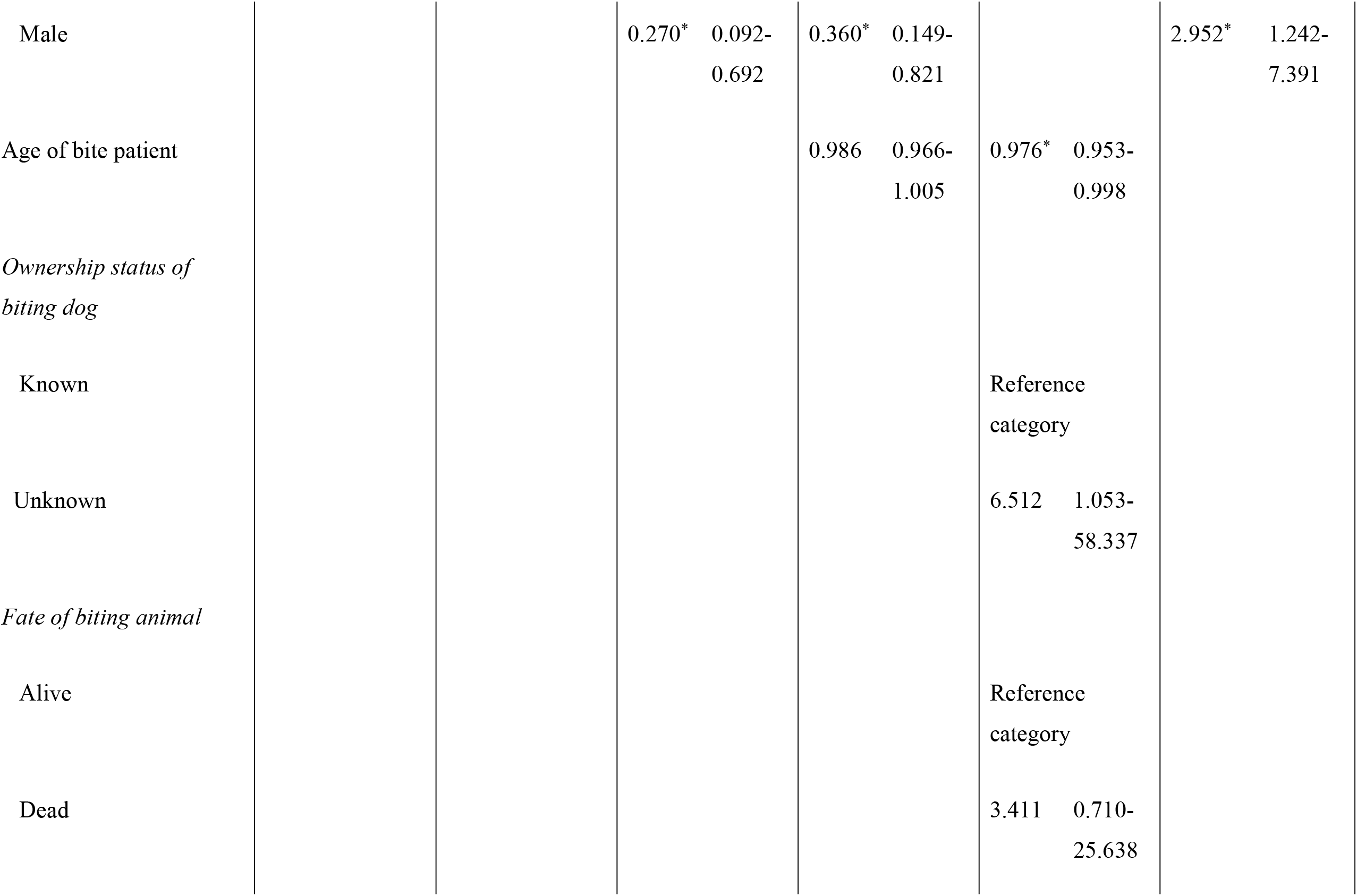
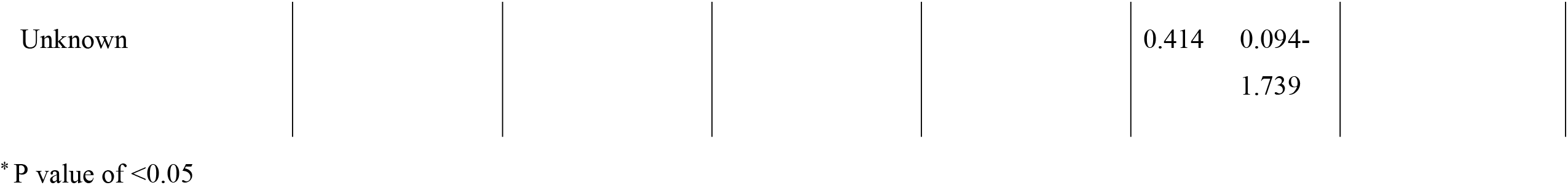
Multivariate analysis of the factors associated with compliance to each of the five doses of PEP, and completion of all the five doses among bite patients in Makueni County.

**Fig 3:**
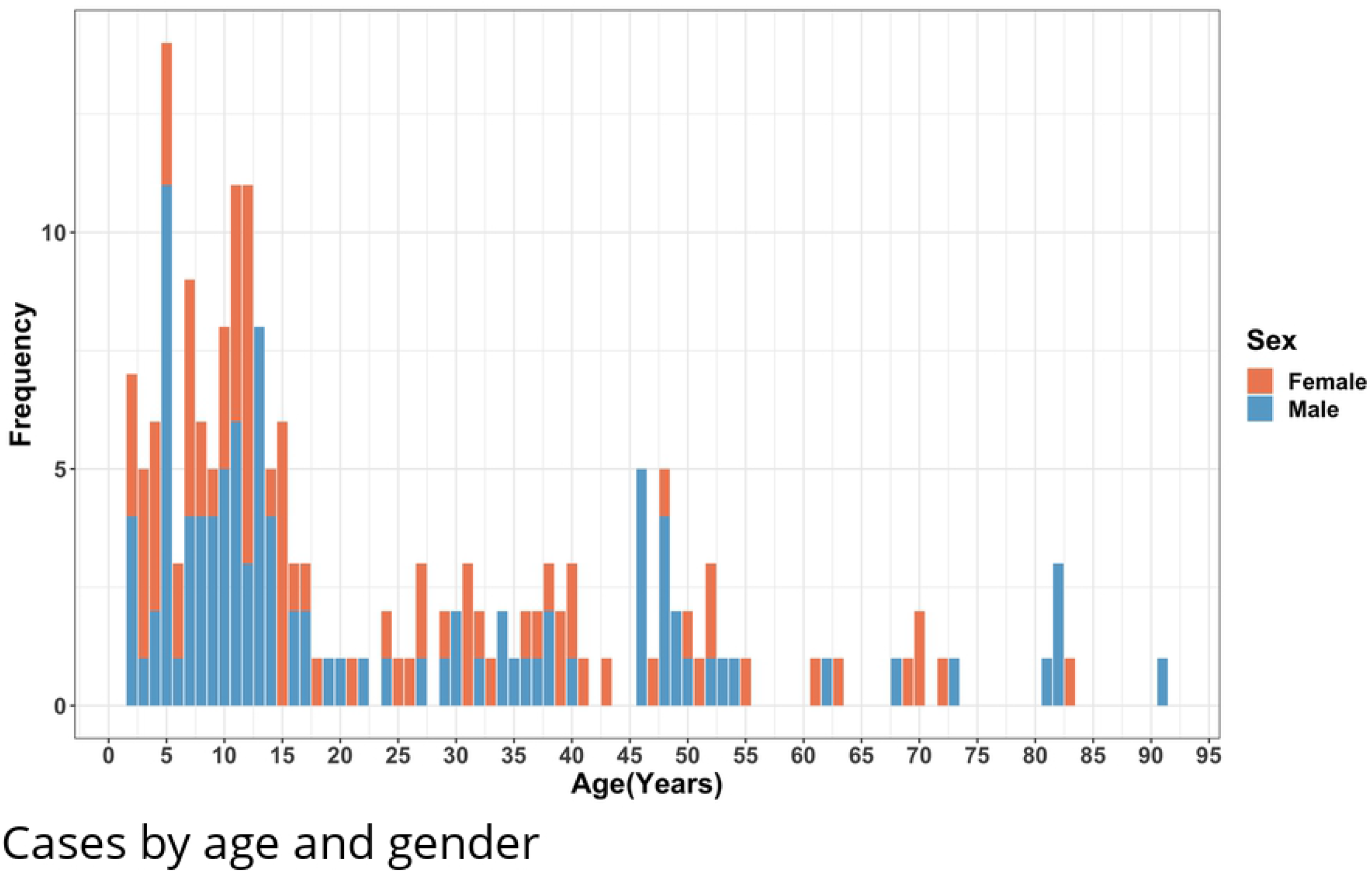
Age distribution of bite patients in years and by gender.

### Factors affecting PEP completion and compliance

Visualisation of the proportion of PEP doses completed per bite category showed that dose two to four had a high uptake of more than 80%. Comparison of PEP dose completion in the intervention and control group showed the intervention group had a relatively higher PEP uptake of more than 70% in all the bite categories while patients in the control group with bite category III had a higher rate of completion of the five doses. The drop-out between vaccinations was higher in the control group especially in the 5^th^ injection where 16 bite patients dropped out (Fig 4).

**Fig 4:**
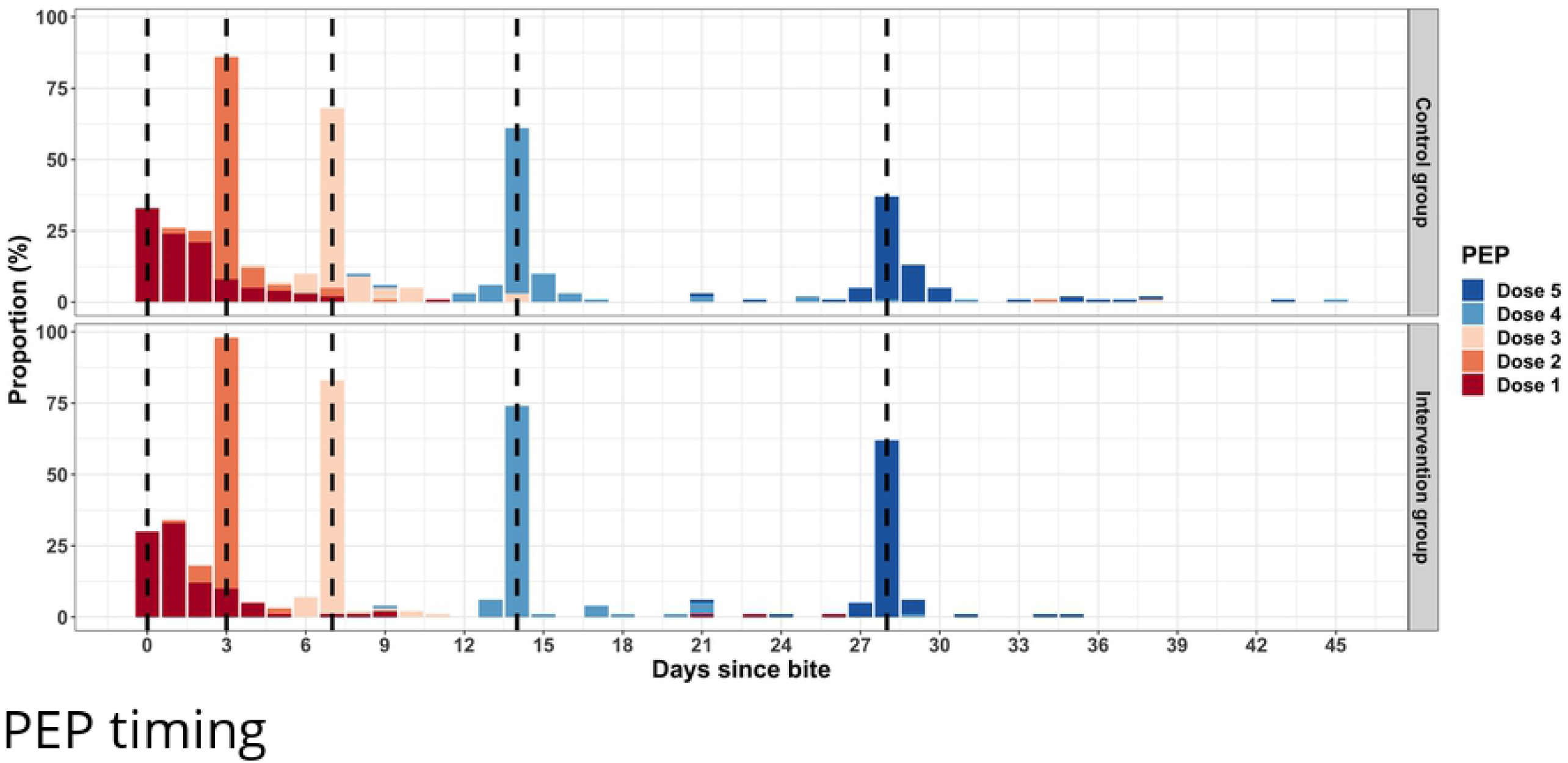
Figure showing the timing of each of the five doses of PEP per the Essen regimen for the control and intervention group. The dashed lines highlight the WHO recommended Essen schedule for the five doses of PEP for bite cases [5].

**Fig 5:**
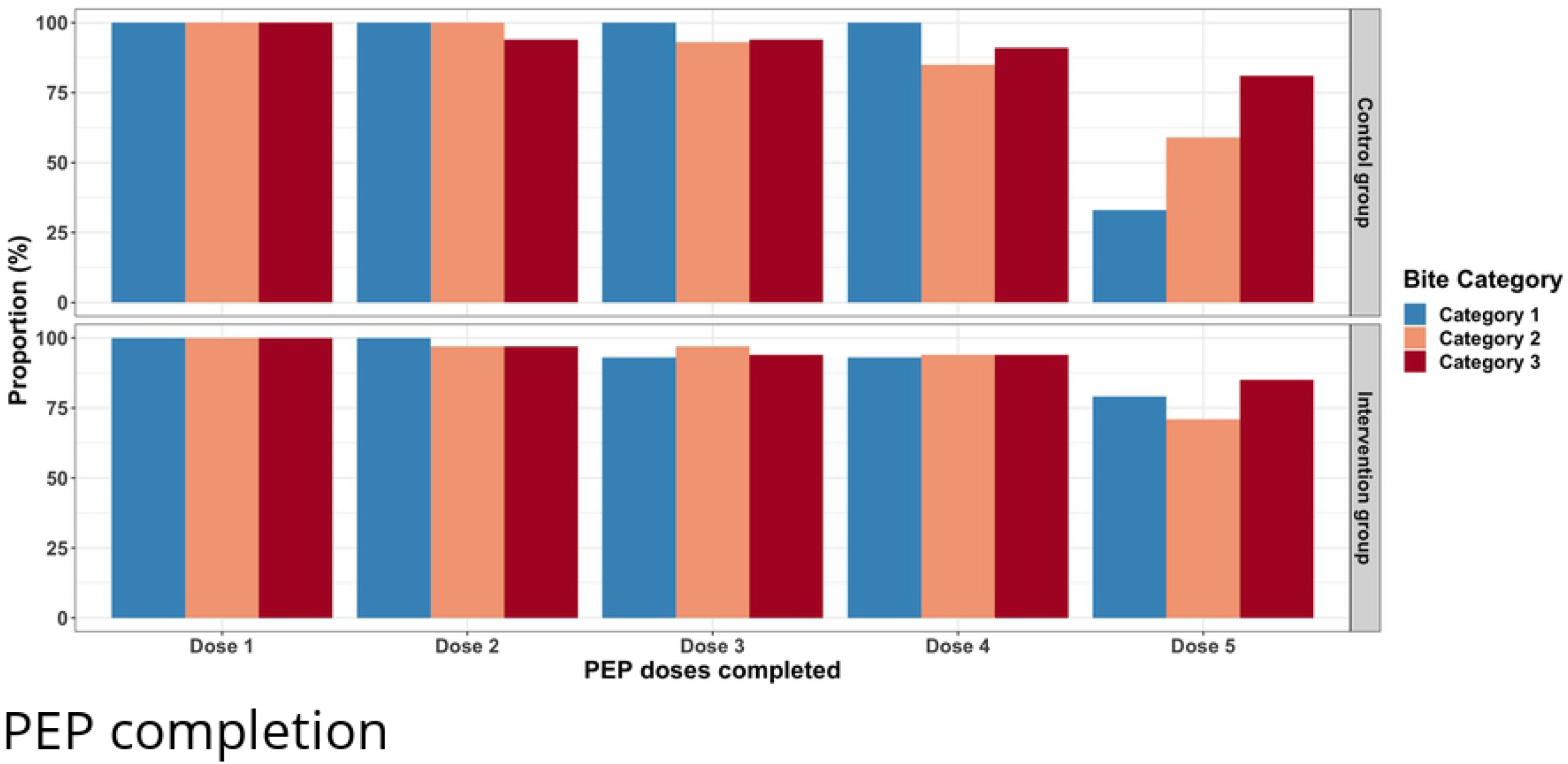
Figure showing the proportion of bite patients who completed the different PEP doses per bite category in the intervention and control group.

In addition, the odds of male patients completing treatment was three times compared to female bite patients (OR 2.95, CI 1.24-7.39). Cost of transport to health facility, monthly family income, age of bite patient, ownership status and fate of biting dog, bite category and home location were not significantly associated with the completion of PEP among the bite patients (Table 3).

Family income per month was significantly associated with the uptake of the first dose less than 24 hours after the bite, where the odds was 3 times (OR 3.2, CI 1.645-6.174) for households with monthly earnings of >USD100.

The odds of compliance with the second dose of PEP due three days after the bite was four times (OR 4, CI 1.33-15.31) in the intervention group, as compared to the control group. This was similar to the uptake of the third to the fifth dose where patients who had been sent SMS reminders had a higher likelihood of complying to the recommended PEP scheduled date (Table 3). Bite patients with category III bites had a higher likelihood of receiving PEP dose 3, 4 and 5 on the recommended scheduled date. In addition, compared to females, male patients had a significantly lower likelihood of PEP compliance during the third (OR 0.3, CI 0.09-0.69) and fourth (OR 0.36, CI 0.15-0.82) dose (Table 3).

A total of 128 (69%) patients were bitten by dogs that were still alive by the time of interview. From this group of patients, 82% (105/128) completed the five doses.

### Reasons for non-compliance and PEP cost

Of the 29 (16%) patients who did not complete the five doses, the main reasons for non-compliance included lack of funds (n=9, 30%), forgetfulness (n=7, 23%) on days for follow-up treatment, unavailability of PEP in the facility (n=5, 17%), that the biting dog was still alive (n=5, 17%), among others (Table 4). However, none of the bite patients in the intervention group failed to complete the five doses due to forgetfulness, while only 22% (n=2/9) that had mentioned lack of funds were in the intervention group. Of those that had mentioned PEP unavailability and that the biting dog was still alive, 60% (n= 3/5) and 40% (n= 2/5) were in the intervention group respectively (Table 4).

**Table 4:**
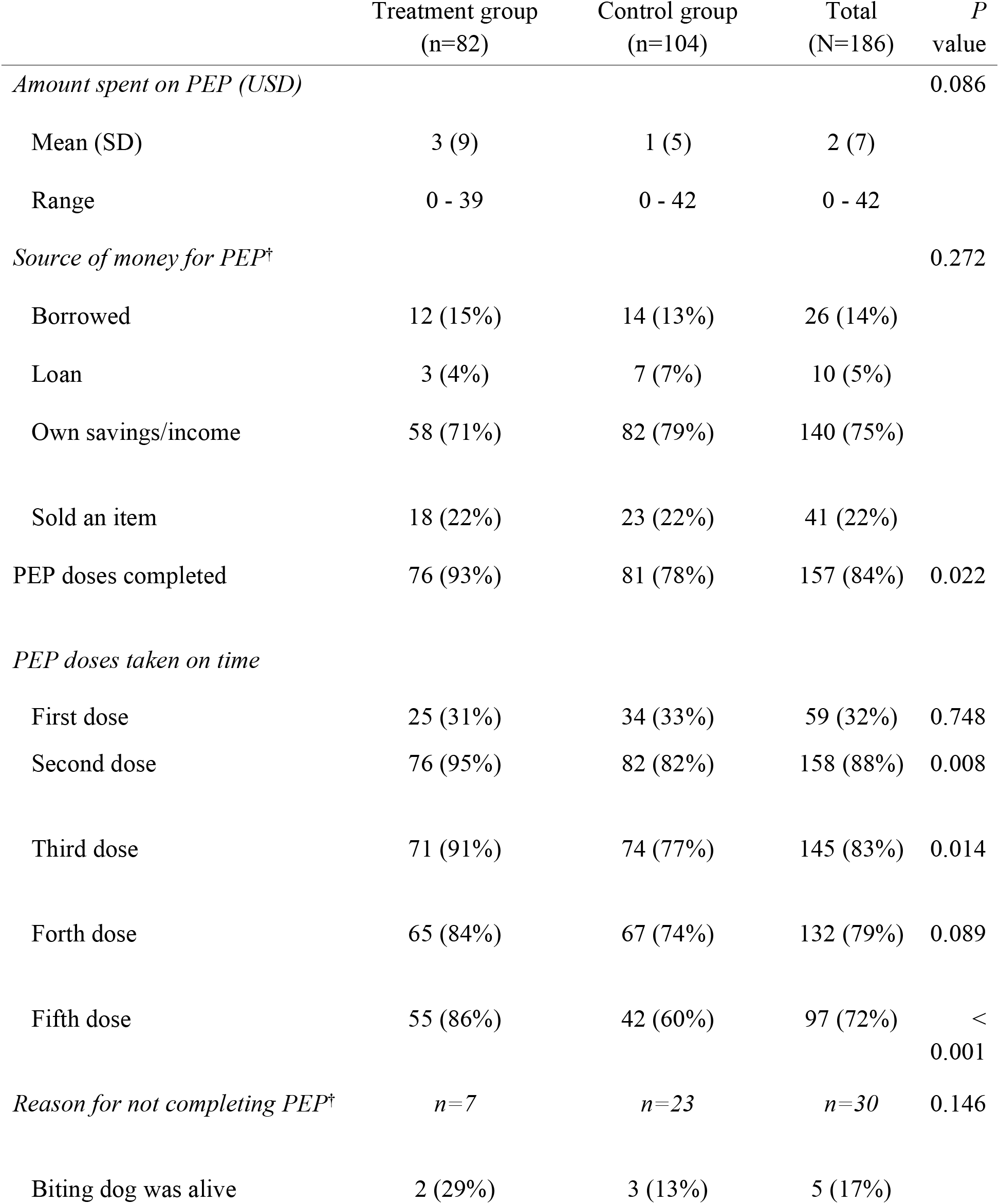
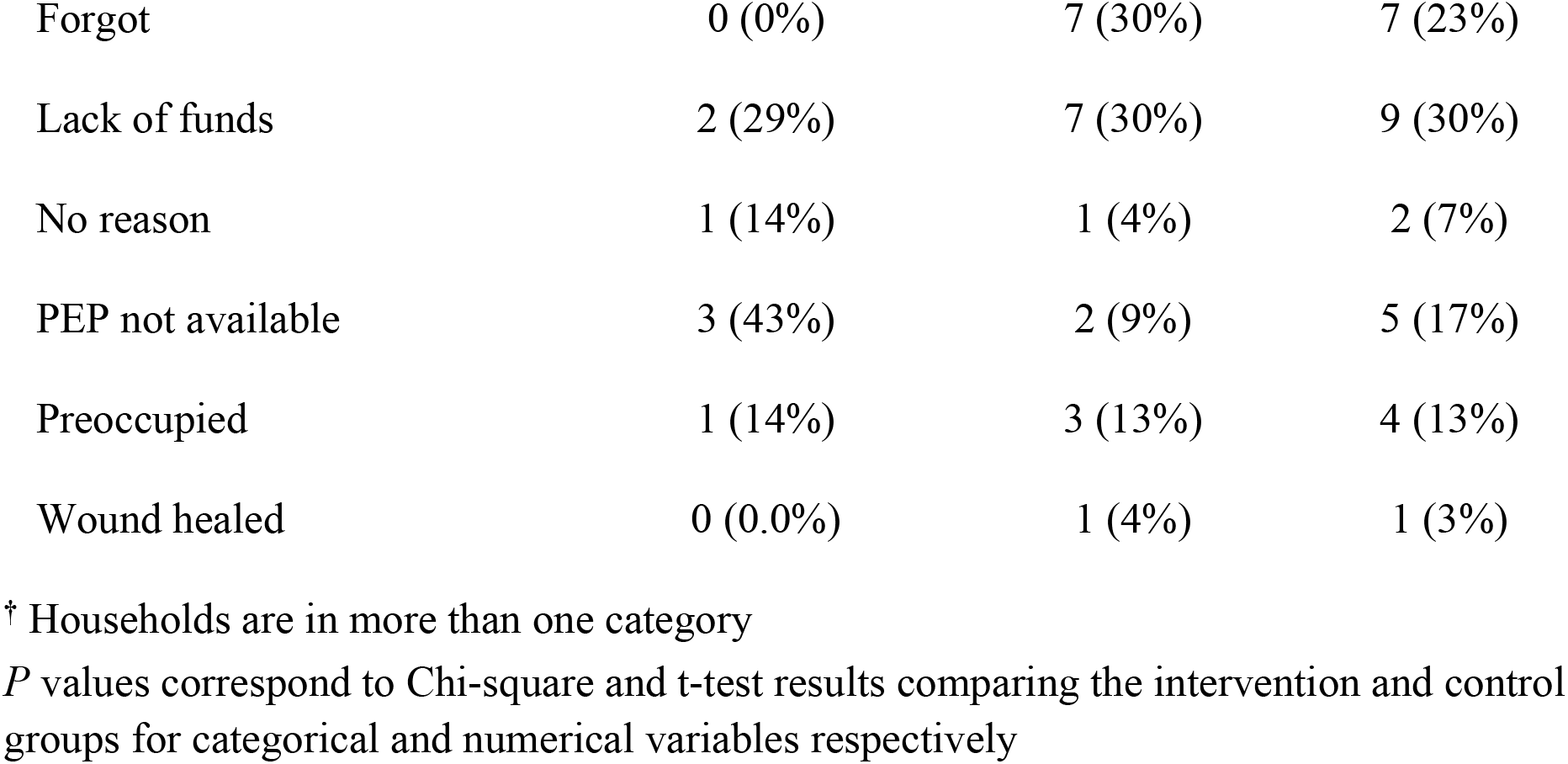
Cost of PEP, completion of PEP doses and reasons for non-completion.

|  | Treatment group<br>(n=82) | Control group<br>(n=104) | Total<br>(N=186) | *P*<br>value |
| --- | --- | --- | --- | --- |
| *Amount spent on PEP (USD)* |  |  |  | 0.086 |
| Mean (SD) | 3 (9) | 1 (5) | 2 (7) |  |
| Range | 0 - 39 | 0 - 42 | 0 - 42 |  |
| *Source of money for PEP<sup>†</sup>* |  |  |  | 0.272 |
| Borrowed | 12 (15%) | 14 (13%) | 26 (14%) |  |
| Loan | 3 (4%) | 7 (7%) | 10 (5%) |  |
| Own savings/income | 58 (71%) | 82 (79%) | 140 (75%) |  |
| Sold an item | 18 (22%) | 23 (22%) | 41 (22%) |  |
| PEP doses completed | 76 (93%) | 81 (78%) | 157 (84%) | 0.022 |
| *PEP doses taken on time* |  |  |  |  |
| First dose | 25 (31%) | 34 (33%) | 59 (32%) | 0.748 |
| Second dose | 76 (95%) | 82 (82%) | 158 (88%) | 0.008 |
| Third dose | 71 (91%) | 74 (77%) | 145 (83%) | 0.014 |
| Forth dose | 65 (84%) | 67 (74%) | 132 (79%) | 0.089 |
| Fifth dose | 55 (86%) | 42 (60%) | 97 (72%) | <<br>0.001 |
| *Reason for not completing PEP<sup>†</sup>* | *n=7* | *n=23* | *n=30* | 0.146 |
| Biting dog was alive | 2 (29%) | 3 (13%) | 5 (17%) |  |

|  |  |  |  |
| --- | --- | --- | --- |
| Forgot | 0 (0%) | 7 (30%) | 7 (23%) |
| Lack of funds | 2 (29%) | 7 (30%) | 9 (30%) |
| No reason | 1 (14%) | 1 (4%) | 2 (7%) |
| PEP not available | 3 (43%) | 2 (9%) | 5 (17%) |
| Preoccupied | 1 (14%) | 3 (13%) | 4 (13%) |
| Wound healed | 0 (0.0%) | 1 (4%) | 1 (3%) |

† Households are in more than one category
*P* values correspond to Chi-square and t-test results comparing the intervention and control groups for categorical and numerical variables respectively

The majority (89%) of the bite patients did not purchase the anti-rabies vaccine since the vaccine was available at the hospital under the Universal Health Coverage program run by the County where healthcare services are free for all households that enroll into the program by paying the USD 5 enrollment fee. Participants who reported incurring costs on PEP spent a minimum of USD 5 and a maximum of USD 45. Nearly all (96%, n=179) of the bite cases incurred transport costs on average of USD 4.6 (range 0 - 45 USD). More than two-thirds of the bite patients (70%) were accompanied to the health facility, resulting in lost earnings and missing school time for school-aged children. In total, 61 (75%) bite cases reported using their own savings to cover treatment costs, while 18 (22%) reported selling livestock (mostly goats and chickens) to cover treatment costs (Table 4).

### Discussion

Here we report on the effect of SMS reminders in increasing proportions of PEP completion among dog bite patients and adherence to the scheduled dates of PEP. We note that SMS reminders were associated with an increased likelihood of completing the PEP doses compared to participants that did not receive SMS reminders. Majority of the patients who did not complete the five doses were in the control group and cited lack of funds and forgetfulness as the main reasons for not completing the doses. In the intervention group, main reported reasons for non completion of the schedule were lack of access to the PEP, lack of funds and biting dog was still alive. The odds of male patients completing treatment was three times compared to female bite patients. Age of bite patient, cost of transport to health facility, monthly family income, ownership status and fate of biting dog, bite category and home location were not significantly associated with the completion of PEP among the bite patients. The intervention group had better compliance on the scheduled doses 2 to 5 with a mean deviation of 0.18 days compared to 0.79 days for the control group. The majority of the bite patients did not purchase the anti-rabies vaccine since the vaccine was available for free at the hospital under the Universal Health Coverage program. Participants that reported incurring cost on PEP spent a minimum of USD 5 and a maximum of USD 45. On average, transportation costs spent by bite patients was USD 4 per visit.

Rabies is a vaccine preventable disease in both humans and animals by annual vaccination of 70% of the dog population to effectively control and eliminate rabies and through prompt administration of PEP to bite patients [2,3,24,25]. Unfortunately, in many countries where rabies is endemic, PEP is unavailable and the cost of vaccine remains high for both individuals and the government, which negatively impacts on the ability of affected countries to meet the Sustainable Development Goals, especially the goal to eradicate extreme poverty and hunger and improve health by 2030 [26,27,28]s. The communities in these endemic countries also remain insufficiently aware of rabies risks and prevention measures. Improving completion and compliance to the PEP regimen is critical. Studies have shown that lack of early and adequate post-exposure vaccination is the most important cause of mortality due to rabies [29,30] with reasons for non-compliance falling under lack of wages, forgotten dates and costs incurred towards treatment [31].

This study highlights the population at risk of contracting rabies as children below the age of 15, likely due their close relationship with dogs. Increasing patient awareness on the risk of contracting diseases, use of SMS reminders and medication subsidies have all been shown to increase compliance with medication [32,33]. However, despite the availability of the Universal Health Coverage program that covers cost for treatment for all members of a household after registration with USD 5, some of the bite patients especially those that did not receive reminders did not complete the five doses of PEP or adhere to the timeliness of each dose of the vaccine. Reasons cited by the bite patients for not completing PEP were forgetfulness and lack of funds. This could be attributed to the inhibitive transport cost to the health facility and cost of PEP for patient not in the Universal Health Coverage program. These results are in accordance to what has been reported for rural areas in Africa where the incidence of rabies is high and the residents are poor and living below the poverty line and have limited access to health care [1, 2]. In addition to SMS reminders, medication subsidies can be used as a synergistic strategy to increase compliance.

Timely access to rabies vaccines by bite patients from suspect rabid dogs and compliance to the full course of the five doses of PEP as recommended by WHO significantly reduces the risk of developing rabies and death. Delays in seeking PEP and incomplete courses exposes bite patients to rabies [6,24]. Here, we see delays in receiving the first dose after the bite and lack of adherence to timeliness on the scheduled day of the subsequent doses of PEP. The delay in seeking PEP and lack of adherence to timeliness to PEP suggests lack of awareness by the community on the need to seek PEP immediately after a bite and adherence to timeliness for the subsequent doses. This study reveals the potential of SMS reminders to improve adherence to PEP schedules as recommended by WHO. Bite patients who received the SMS reminders had a higher likelihood of complying with timeliness of the subsequent doses after the initial dose, potentially both reminding the patient and reinforcing their awareness about the need for complete and timely PEP.

As many governments express interest in mobile health as a complementary strategy for strengthening health systems and achieving the health-related Sustainable Development Goals in low and middle income countries, the potential of mobile phone technology in improving health has been shown in different health sectors [11]. This includes use of mobile phones in rabies surveillance and demonstration of PEP demand in health facilities [34]. In delivery of health interventions, SMS has been shown to increase completion of the five doses of PEP (irrespective of age or location of the bite patient) [35] and to improve health outcomes, post-treatment hospital return and adherence to health diets and medication especially in HIV/AIDS and diabetes patients with up to 100% effectiveness [14,18,37,38]. In our study, the odds of completion of the five doses was 3.4 times in patients who received the SMS reminder compared to those who did not receive the SMS reminder.The effect of the SMS reminder was irrespective of the age of the bitten patient, age of the household head, fate of the biting dog, occupation of the bite patient/next of kin, category of bite, vaccination status of the biting dog, education level, ownership of health cover, family monthly income, livestock ownership status, transport cost, distance to the health facility, source of money used for treatment and home location (urban/rural) of the bite patient.

Although the study shows significantly improved completion rates of PEP by bite patients who received SMS reminders, the study had a limitation in that data were collected at different times of the year. This could result in a bias to compliance for PEP as time and availability of resource may have not been favorable at that specific time. The design of the study also lacked randomization of the patients into control and intervention group due to ethical consideration. The use of the SMS reminder strategy may also not be effective for older patients with lower literacy levels, lower socioeconomic status or communities without the power to purchase a phone and in rural areas where network connection is poor.

## Conclusion

Rabies continues to pose a significant public health concern in Kenya. Prevention of clinical cases of human rabies following exposure is dependent on prompt access and compliance to PEP regimens. This study highlights the potential value of text messages in delivery of public health interventions as a complementary strategy for strengthening health systems. Integrating SMS as a reminder on the next dose of PEP to dog bite patients at risk of contracting rabies should work synergistically with efforts to control and eliminate rabies in endemic countries including provisioning of PEP supported by Gavi, the Vaccine Alliance. A high level of community awareness on the need to seek PEP immediately after a bite and adhere to PEP schedule is critical to reduce the number of human rabies cases and achieve the ‘Zero by 30’ goal.

## Data Availability

The dataset generated and analyzed for this study is available from Open Science Framework: https://osf.io/zxeqr/. This dataset is available under a CC0 1.0 Universal license and DOI - DOI 10.17605/OSF.IO/ZXEQR

## Acknowledgement

We appreciate dog bite patients from the Makueni County Referral Hospital for their participation in the study.

## Contributorship

***Veronicah Mbaire Chuchu:*** Conceptualization, Investigation, Data Curation, Formal Analysis, Methodology, Supervision, Writing – Original Draft Preparation, Writing –Review & Editing; **Mutono Nyamai:** Data Curation, Formal Analysis, Writing – Review & Editing; **Bichanga Philet:** Methodology, Writing – Review & Editing; **Kitala Philip**: Supervision, Writing – Review & Editing; **Ksee Daniel:** Writing – Review & Editing; **Muturi Mathew:** Writing – Review & Editing; **Mwatondo Athman**: Writing – Review & Editing; **Nasimiyu Carolyne**: Writing – Review & Editing; **Lawrence Akunga:** Writing – Review & Editing; **Amine Amiche:** Writing – Review & Editing**; Katie Hampson:** Conceptualization, Formal Analysis, Funding Acquisition, Methodology, Resources, Writing – Review & Editing; **SM Thumbi:** Conceptualization, Funding Acquisition, Project Administration, Resources, Supervision, Writing Review & Editing

**S1 CONSORT checklist for Effect of phone text message reminders on compliance with rabies post-exposure prophylaxis following dog-bites in rural Kenya**.

**S2 Appendix: A questionnaire on factors associated with completion to the five doses of anti-rabies vaccine in Makueni County**.

**S3 Clinical trial protocol**

